# Efficacy and Safety of CKD-495 in the Treatment of Gastritis: A Systematic Review and Meta-analysis

**DOI:** 10.1101/2025.07.18.25331707

**Authors:** Saad Arsalan Wasti, Mahnoor Arfan, Omar Abdullah Gill, Meeram Noor, Maryyam Aqeel, Hammad Javed

**Author notes:** **Corresponding Author:** Name: Mahnoor Arfan, Address: 141 D Block, Izmir Town, Lahore, Pakistan. **Funding source:** None.

## Abstract

**Background:** Gastritis is a common condition affecting nearly half the world’s population. The Standard treatments like proton pump inhibitors have limitations such as adverse effects, frequent relapses, and growing antibiotic resistance in H. pylori. New therapies such as CKD-495 could offer safer and more effective alternatives by addressing these challenges.

**Data Sources:** We conducted a systematic review and meta-analysis following PRISMA guidelines. Detailed Searches were performed in PubMed, Scopus, Embase, and ClinicalTrials.gov up to April 2025. Additional studies were identified through bibliographic mining and citation searches.

**Main Findings and Limitations:** Two randomized controlled trials (total N=475) that compare CKD-495 with Artemisiae argyi folium (AAF) met inclusion criteria. The meta-analysis showed that CKD-495 significantly improved rates of erosion healing (OR≈2.1; P=0.001) compared to AAF. There was no increase in adverse events. The risk of bias was low across all domains. limitations include the small number of studies, limited sample sizes, and short follow-up periods restrict the generalizability of the findings.

**Conclusions:** CKD-495 appears to be a promising alternative for treating gastritis. this study demonstrates the better mucosal healing with similar safety compared to AAF. Further large, multicenter trials are needed to confirm these results and evaluate long-term outcomes.

## FULL TEXT

Gastritis, an inflammation of the stomach lining, presents with various gastrointestinal symptoms such as bloating, nausea, epigastric pain, and reduced appetite (1). Common causes of gastritis include *H. pylori* infection, drugs, bile reflux and autoimmunity (2). Gastritis is a widespread condition; affecting ~50% of the world’s population (3). Currently, proton pump inhibitors are the standard for gastritis treatment. However, they may cause gastric gland toxicity (4). The pathophysiology of chronic gastritis is complex. Western medicine targets *H. pylori*, but often neglects the contributing host factors. Thus, relapses occur frequently (5,6). Antibiotic resistance of *H. pylori* has also developed significantly. This may predispose to severe reinfections (6,7). Despite the muco-protective and acid-release inhibiting effects of PPIs, the disease cannot be fully eradicated. Moreover, these drugs have various adverse effects (5,8). A possible alternative is traditional Chinese medicine which has shown promising results in eliminating *H. pylori* by enhanced bactericidal properties (7). A Phase III superiority clinical trial has demonstrated that CKD-495, a South Korean-origin medicine for gastritis, improved treatment outcomes with better erosion rates (8). It acts as an anti-inflammatory agent and also protects the gastric mucosa. This systematic review and meta-analysis compares CKD-495’s efficacy and safety against *Artemisiae argyi folium* (AAF) in gastritis patients.

We conducted this systematic review and meta-analysis in line with the Preferred Reporting Items for Systematic Reviews and Meta-Analyses (PRISMA) guideline. A literature search was conducted on PubMed, Scopus, Embase and ClinicalTrials.gov till April 2025. The complete search strings are provided in the supplementary **Suppl1**. Bibliographic mining and citation searching were also done. The records obtained were uploaded to *Rayyan®* for screening. Duplicates were resolved, and the remaining records were screened independently by two reviewers. Conflicts were resolved through mutual consensus or through a third reviewer (see **Suppl1** for the detailed criteria used). Data extraction was done independently by two reviewers on an *Excel*® spreadsheet, for base-line characteristics and for the outcomes: ‘Improvement Rate of Erosion’ and ‘Adverse Events’. Disagreements were resolved through a senior reviewer. Risk of bias for the included RCTs was assessed independently by two reviewers, using the Cochrane Risk of Bias Tool RoB 2.0 across five domains (9). *Meta* and *metafor* packages in R software (version 4.4.3) were used for meta-analysis. Outcomes were analysed by the MH formula, with a random-effects model. Dichotomous outcomes were pooled into odds-ratios (OR) with 95% confidence intervals (CI) [p < 0.05 was considered statistically significant]. *Tau*^*2*^, *Chi*^*2*^, *df* and *I*^*2*^ statistics were used to investigate heterogeneity among studies, with the overall effect being measured by *Z*-statistics. Publication bias was not assessed due to limited studies.

The literature search identified 78 studies, of which 37 were removed as duplicates. The remaining 41 studies were assessed on title-abstract, leading to exclusion of 38 studies. Full texts of the remaining 3 studies were assessed for eligibility. One study, from citation searching, was also assessed. A total of 2 studies met the criteria and were included in the analysis. The study selection process is illustrated in detail in **Suppl1**. The risk of bias in the included studies, assessed using the RoB 2.0 tool, revealed an overall ‘low’ risk of bias across all five domains, indicating high quality and consistency among studies. The findings were visually represented as the traffic light and summary plots.

Two RCTs were included with a total of 475 patients, having a mean age of 47.16±14.55 years, of which 32.75% were males. Gastrointestinal symptom scores averaged from 4.6-5.5. Acute gastritis cases ranged from 13-61 patients, whilst chronic gastritis ranged from 34-53. Baseline endoscopy showed mostly Grade 1–2 erosions, edema, redness, and hemorrhage across acute and chronic gastritis groups, indicating comparable clinical and endoscopic profiles. Detailed study characteristics and patient demographics are presented in **Table 1**.

**Table 1.**
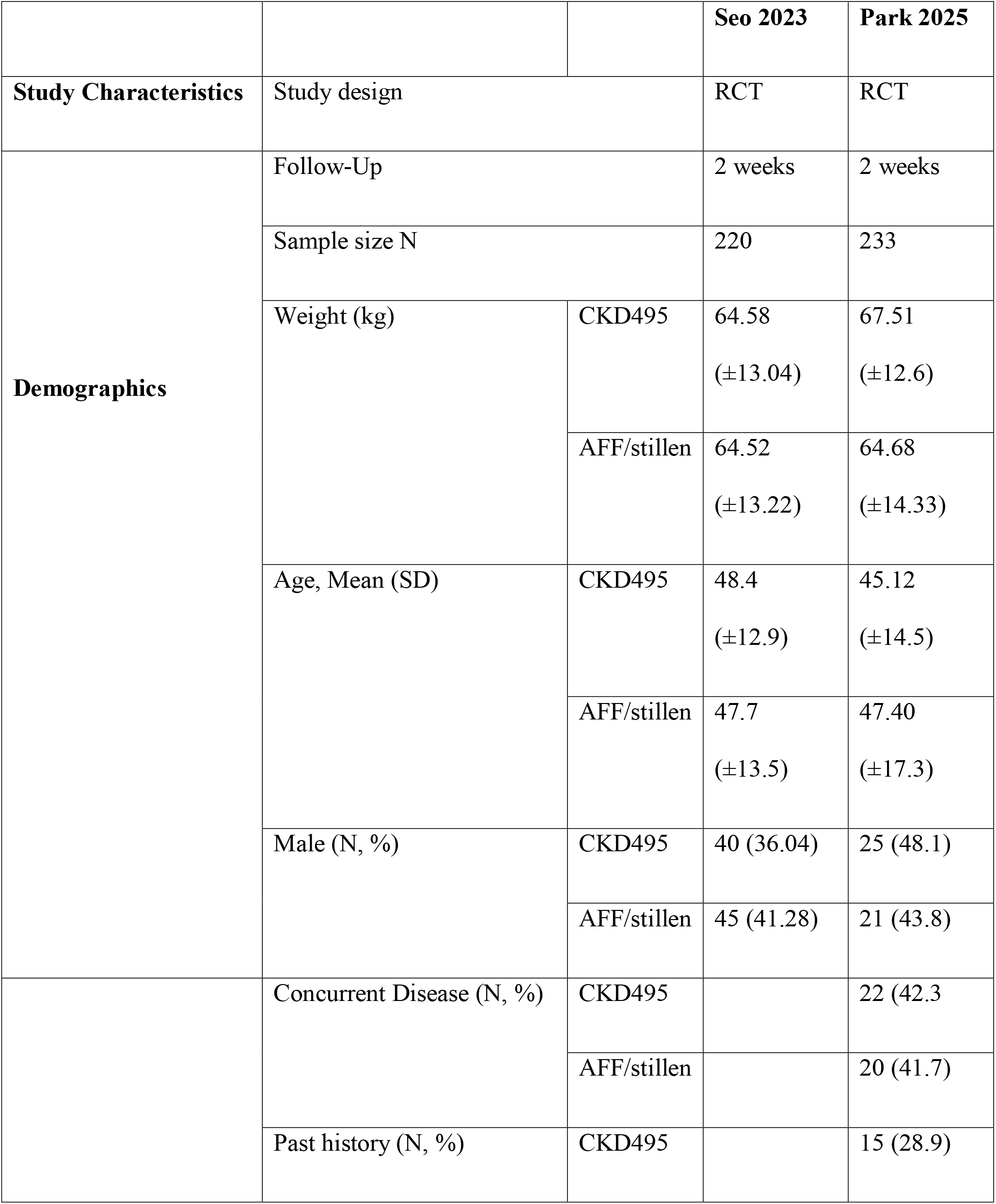
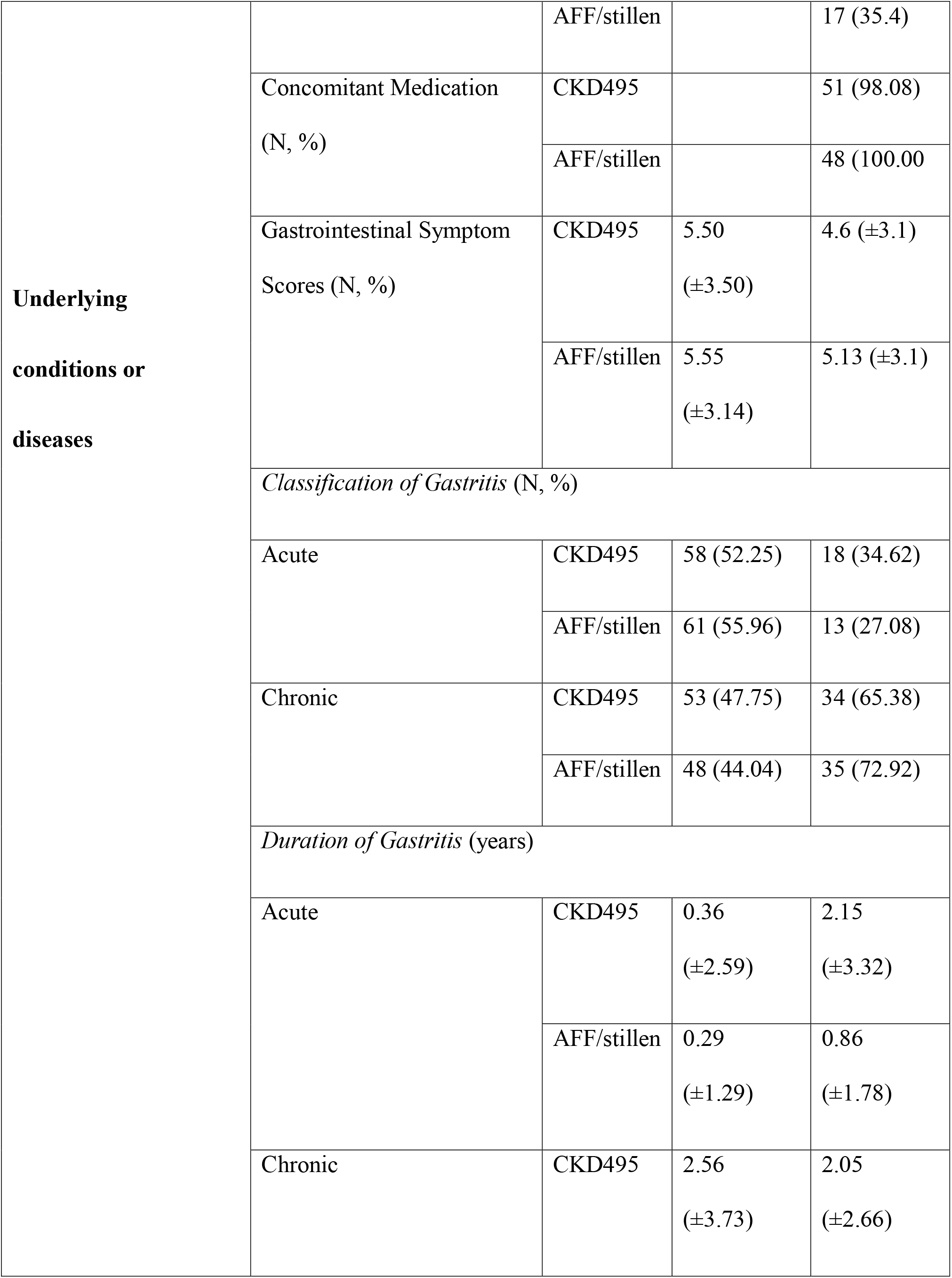
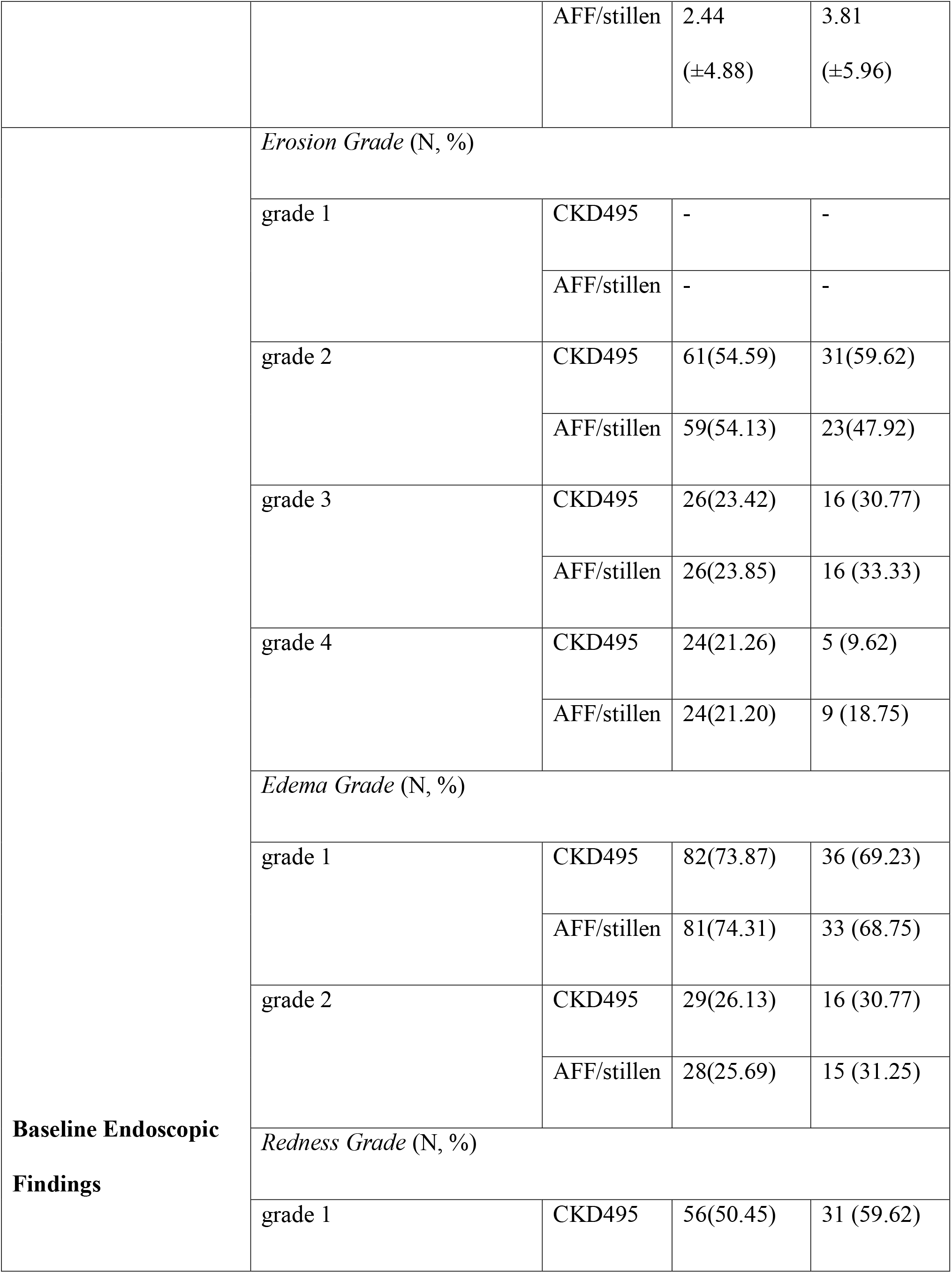
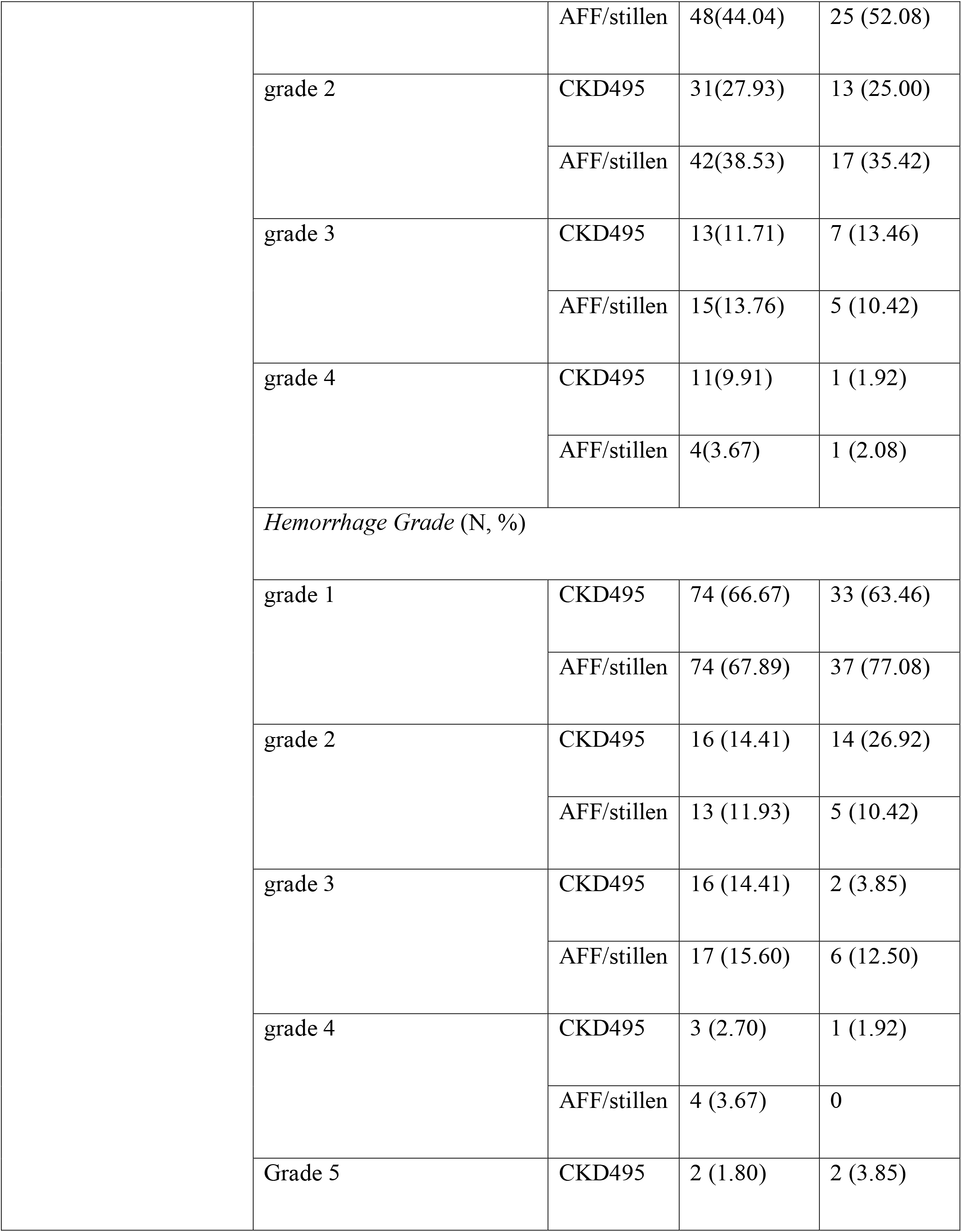
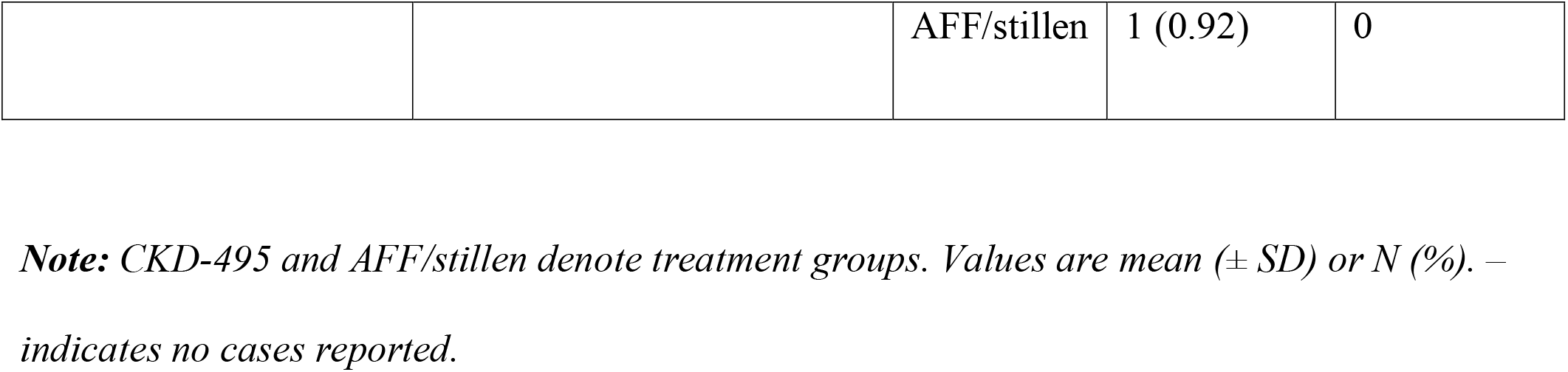
Baseline Characteristics of Included Studies.

All included studies reported incidence of any adverse events per Safety Set (SS, N=344; CKD-495: 176, AAF: 168), with *no statistically significant* difference in the incidence of experiencing any AE between the CKD−495 and AAF groups (OR=0.82, 95% CI 0.29-2.32, P=0.707). Inter-study heterogeneity was negligible (I^2^=0%). In the Full Analysis Set (FAS, N=320; CKD-495:157, AAF:163), CKD-495 significantly outperformed AAF (OR=2.09, 95% CI 1.33-3.28, P=0.001; I^2^=0%) with regards to improvement in rate of erosion. Similar results were observed in the Per Protocol Set (PPS, N=301; CKD-495:145, AAF:156) with a comparable effect size (OR=2.17, 95% CI 1.36-3.46, P=0.001; I^2^=0%).

This meta-analysis compared the efficacy and safety of CKD-495 to an active control (AAF, also called Stillen), in patients having gastritis. The analysis of the pooled data from the studies showed that participants who received CKD-495 had a significantly better rate of erosion healing compared to the control group, in both the FAS and PPS. There are no other differences concerning the occurrence of adverse events. This data thus demonstrates the therapeutic efficacy of CKD-495 as a safe and effective treatment for gastric mucosal damage, compared to AAF. Even though these findings are still preliminary, they strongly suggest the promising nature of CKD-495.

To ensure proper reproducibility and systematic inclusions, we followed PRISMA guidelines. The low risk of bias significantly strengthens the pooled results. Clinically validated tools such as FAS and PPS are implemented to show treatment effects. Endoscopic grading of gastric lesions was used to objectively assess improvement regarding mucosal healing. OR were used to compare the likelihood of mucosal healing between CKD-495 and AAF groups, while statistical significance was evaluated using P-values. The I^2^ statistic was negligible for all analyses, indicating exceptional consistency among included trials. This indicates that CKD-495’s favourable effects were consistent and not influenced by differences in study design, patient population, or how outcomes were assessed.

This is the earliest meta-analysis that studies CKD-495’s efficacy and safety in gastritis. Although this study effectively assessed CKD-495’s role in gastritis patients, it has its limitations. Long term outcomes like recurrence rates and persistent mucosal healing require validation with larger multi-centered RCTs with extended follow-up periods. It is also essential to highlight that the limited number of studies and small sample sizes constrain generalisability of these findings to wider patient populations. Furthermore, *H. pylori*-related, NSAID-induced, or stress-gastritis subtype stratified analyses could add additional explanatory power into the treatment effects on patients. In conclusion, CKD-495 is a promising candidate in gastritis treatment, though more research is needed to validate these findings.

## Data Availability

All data produced in the present study are available upon reasonable request to the authors

## Author Declarations

**CRediT Authorship Contribution Statement**

**S.A.W**.: Conceptualisation, Methodology, Software, Validation, Formal Analysis, Investigation, Resources, Data Curation, Writing – Original Draft, Writing – Review & Editing, Visualisation, Supervision, Project Administration; **M.Ar**.: Validation, Methodology, Software, Investigation, Writing – Original Draft, Writing – Review & Editing, Visualisation, Supervision, Project Administration; **O.A.G**.: Conceptualisation, Methodology, Software, Validation, Formal Analysis, Data Curation, Writing – Review & Editing, Visualisation, Supervision; **M.N**.: Methodology, Software, Validation, Investigation, Writing – Original Draft, Visualisation, Supervision; **M.Aq**.: Methodology, Investigation, Writing – Original Draft; **H.J**.: Validation, Writing – Original Draft, Writing – Review & Editing.

## List of Supplemental Digital Content

Supplemental Digital Content 1. PDF

Suppl1. Supplementary Materials file including

- **Supplementary Table 1**. Detailed Database Search Strings
- **Supplementary Table 2**. Inclusion and Exclusion Criteria
- **Supplementary Figure 1**. Risk of Bias Summary Plot
- **Supplementary Figure 2**. Risk of Bias Traffic Light Plot
- **Supplementary Figure 3**. PRISMA Flowchart

