## Supplementary Material for "Efficacy and Safety of CKD-495 in the Treatment of Gastritis: A Systematic Review and Meta-analysis"

**Supplementary Materials**

**Supplementary Table 1.** Databases and Search String (Search Date: 14/04/2025)

| **Databases** | **Search String** | **Results** |
| --- | --- | --- |
| ***PubMed*** | #1  "gastritis"[MeSH Terms] OR "Gastritides"[Title/Abstract] OR "gastric inflammation"[Title/Abstract] OR "gastric inflammatory disease"[Title/Abstract] OR ("gastritic"[All Fields] AND "disease"[Title/Abstract]) OR ("gastritic"[All Fields] AND "disorder"[Title/Abstract]) OR "gastritis"[Title/Abstract] OR "granulomatous gastritis"[Title/Abstract] OR "reflux gastritis"[Title/Abstract] OR "stomach inflammation"[Title/Abstract]  #2  "ckd-495"[Title/Abstract] OR "ckd495"[Title/Abstract] OR "ckd-495"[Title/Abstract] OR "cinnamomum cassia"[Title/Abstract] OR "cinnamomum aromaticum"[Title/Abstract]  #3  #1 AND #2 | **15** |
| ***Embase*** | ('gastritis'/exp OR 'biliary gastritis' OR 'caustic gastritis' OR 'gastric inflammation' OR 'gastric inflammatory disease' OR 'gastritic disease' OR 'gastritic disorder' OR 'gastritides' OR 'gastritis' OR 'granulomatous gastritis' OR 'reflux gastritis' OR 'stomach inflammation') AND ('ckd-495' OR 'ckd495' OR 'ckd 495' OR 'cinnamomum cassia'/exp OR 'cinnamomum aromaticum' OR 'cinnamomum cassia') | **23** |
| ***Cochrane*** | ID Search Hits  #1 MeSH descriptor: [Gastritis] explode all trees 858  #2 (gastritis):ti,ab,kw 3141  #3 (Gastritides):ti,ab,kw 0  #4 (gastric inflammat*):ti,ab,kw 2053  #5 (gastritic dis*):ti,ab,kw 2  #6 (granulomatous gastritis):ti,ab,kw 0  #7 (reflux gastritis):ti,ab,kw 327  #8 (stomach inflammat*):ti,ab,kw 1780  #9 #1 OR #2 OR #3 OR #4 OR #5 OR #6 OR #7 OR #8 5735  #10 (ckd-495):ti,ab,kw 4  #11 (ckd495):ti,ab,kw 0  #12 (cinnamomum cassia):ti,ab,kw 51  #13 ("c. cassia"):ti,ab,kw 5  #14 (cinnamomum aromaticum):ti,ab,kw 32  #15 ("c. aromaticum"):ti,ab,kw 4  #16 #10 OR #11 OR #12 OR #13 OR #14 OR #15 68  #17 #9 AND #16 4 | **3** |
| ***Scopus*** | #1  TITLE-ABS-KEY(gastritis) OR TITLE-ABS-KEY(Gastritides) OR TITLE-ABS-KEY(gastric inflammat*) OR TITLE-ABS-KEY(gastritic dis*) OR TITLE-ABS-KEY(granulomatous gastritis) OR TITLE-ABS-KEY(reflux gastritis) OR TITLE-ABS-KEY(stomach inflammat*)  #2  TITLE-ABS-KEY(ckd-495) OR TITLE-ABS-KEY(ckd495) OR TITLE-ABS-KEY(ckd 495) OR TITLE-ABS-KEY(cinnamomum cassia) OR TITLE-ABS-KEY(cinnamomum aromaticum)  #3  #1 AND #2 | **37** |
| ***Bibliographic Mining And Citation Searching*** | - | **1** |

**Supplementary Table 2.** Inclusion and Exclusion Criteria

|  | **Inclusion** | **Exclusion** |
| --- | --- | --- |
| **Population** | Adults diagnosed with gastritis | Children  Adults with any condition other than gastritis  Animal studies  Healthy adults |
| **Intervention** | CKD-495 | Any other intervention |
| **Comparator** | Artemisiae argyi folium | Any other comparator |

**Supplementary Figure 1. Risk of Bias Assessment (Traffic Plot)**

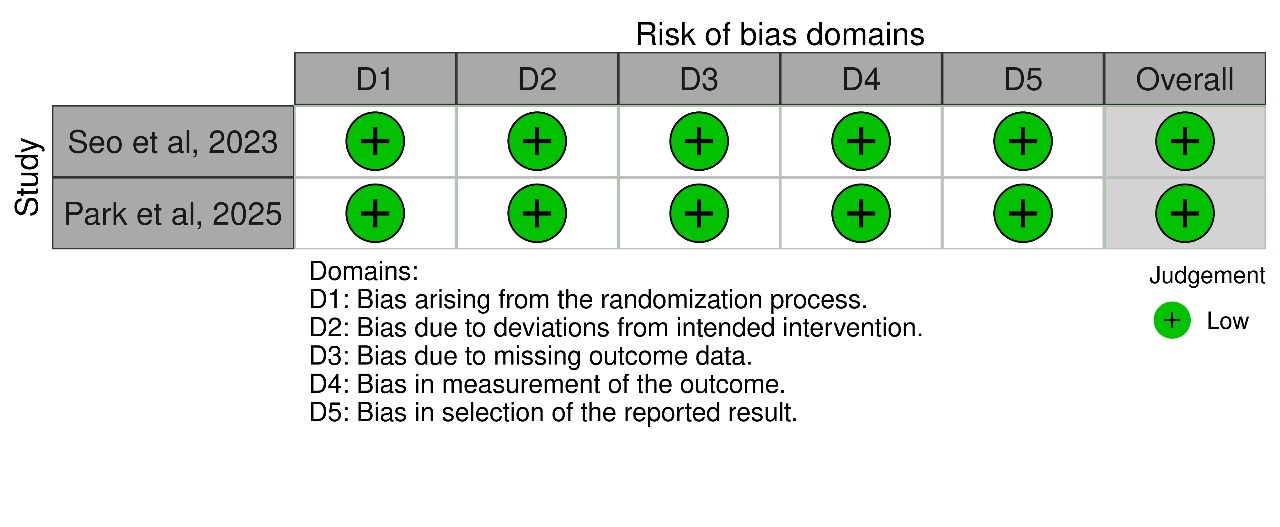

**Supplementary Figure 2. Risk of Bias Assessment (Summary Plot)**

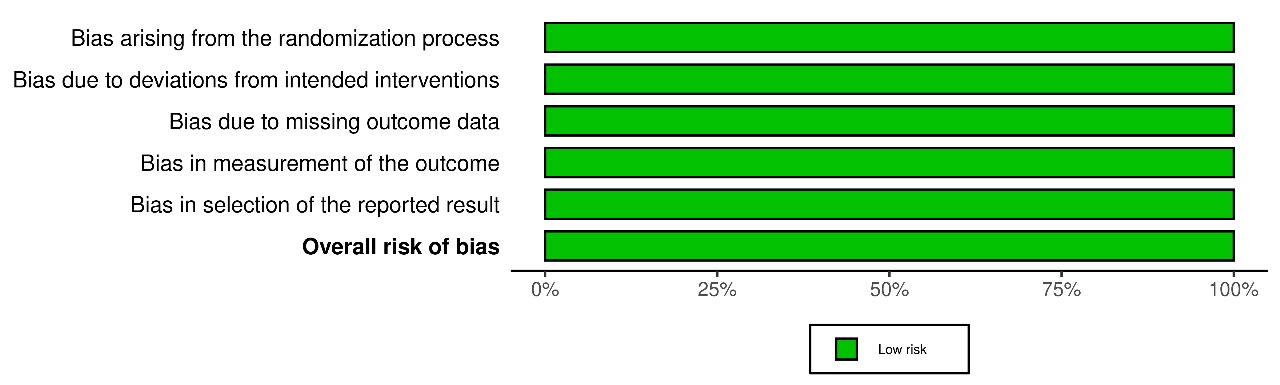

**Supplementary Figure 3. PRISMA flowchart**

**CKD-495 Figure 1**

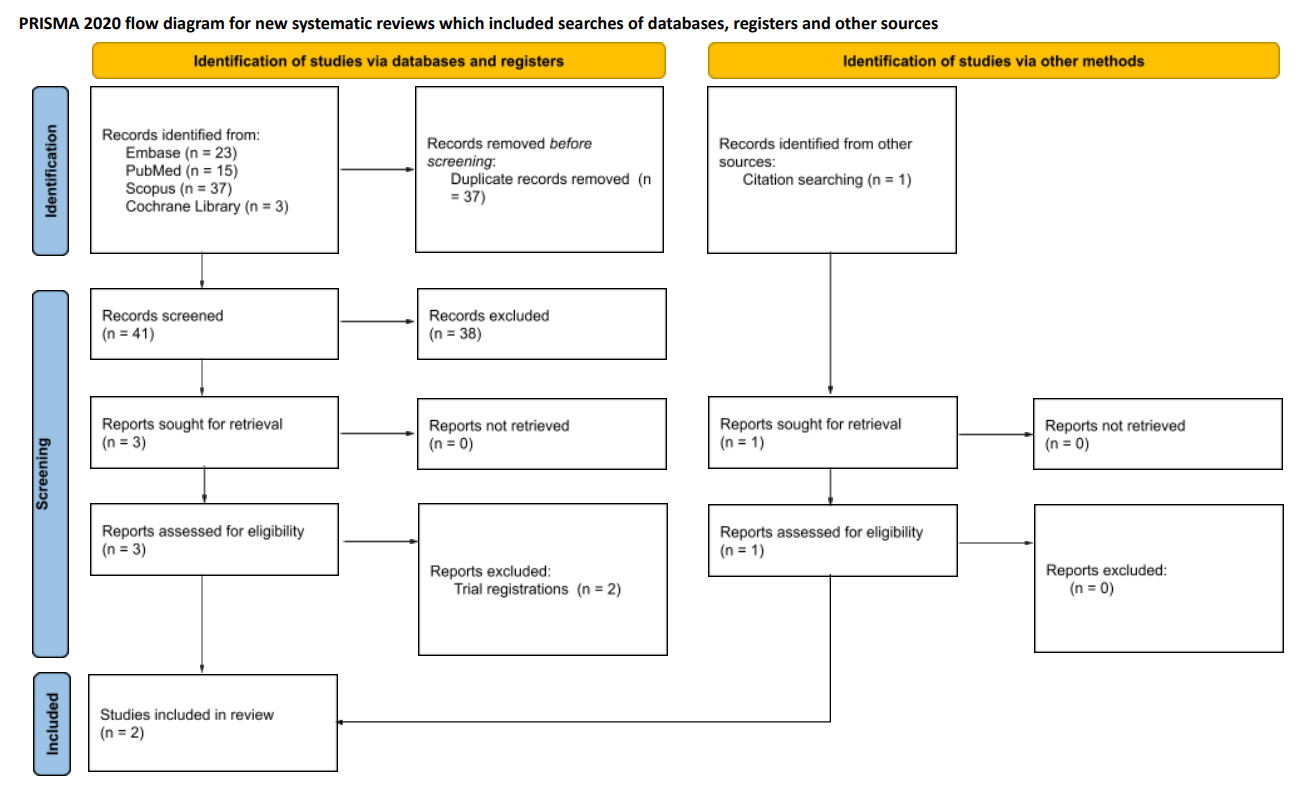
